# AlloGraph: open-source web-based scalable platform for registry-driven monitoring of allogeneic hematopoietic cell transplant activity

**DOI:** 10.64898/2026.06.29.26354738

**Authors:** Axel Cousin, Vivien Legrand, Raynier Devillier, Micheline Karam, Edouard Forcade, Charlotte Jubert, Alban Villate, Martin Eloit, Emmanuel Gyan, Patrice Chevallier, Hélène Labussière-Wallet, Cristina Castilla-Llorente, Johan Maertens, Patrice Ceballos, Marie-Thérèse Rubio, Bénédicte Bruno, Yves Chalandon, Xavier Poiré, Jean-Baptiste Méar, Virginie Gandemer, Jacob Levy, Florent Malard, Philippe Lewalle, Catherine Paillard, Michael Loschi, Jean-Hugues Dalle, Amandine Charbonnier, Etienne Daguindau, Jacques-Olivier Bay, Pedro H. Prata, Natacha Maillard, Felipe Suarez, Aline Schmidt, Malek Benakli, Ali Bazarbachi, Julian Thalhammer, Stephanie Nguyen, Nicole Raus, Anne Huynh, David Michonneau, Nicolas Vallet, the SFGM-TC

## Abstract

Despite longitudinal and multidimensional collected data within registries, their routine exploitation for value-based care and outcome transparency remains limited by analytical complexity and heterogeneous expertise across centers. To address this gap, we developed an open-source and free web-based software which allows registry-based data analysis operational for evaluation of practices and quality system management applied to allogeneic hematopoietic cell transplant registry. It was built with Python and Dash framework to treat user formatted data. AlloGraph produces epidemiological summaries, survival analyses, and quality management indicators. Privacy protection is ensured by a Transport Layer Security protocol to a secure server where processing occurs in-memory, without data saving. AlloGraph was evaluated positively by 30 practitioners in 24 transplant centers, of whom 89% anticipated that AlloGraph would change their monitoring practice. AlloGraph represents a privacy-preserving and user-centered platform simplifying registry analysis for activity monitoring. This scalable model could be adapted to exploit real-world health databases.

## INTRODUCTION

Clinical dashboards allow for an automated and rapid analysis and visualization of complex clinical datasets. The use of dashboards is associated with improved adherence to quality guidelines and might support clinical decision.^1,2^ In the setting of intensive care, a randomized study revealed that, compared to routine tools, the use of a dashboard was associated with shorter time to collect data (10.4 vs. 4 min) and higher frequency of adequately represented data.^3^

Allogeneic hematopoietic cell transplantation (allo-HCT) is a major treatment for malignant hematological diseases, notably acute myeloid and lymphoid leukemias. Aplastic anemia and inborn errors are the first indications for non-malignant diseases.^4^ Depending on the context, the objectives of allo-HCT can be: (i) to replace recipient hematopoietic cells by those of the donor, (ii) to correct defective functions in patients transplanted for metabolic disorders, or (iii) in the context of hematological malignancies, to provide donor immune cells with an antitumor activity, also known as graft-versus-leukemia (GVL) effect.^5^ These immune cells may also react against heathy tissues, initiating an acute or chronic graft-versus-host disease (a/c GVHD).^6,7^ The main causes of mortality are disease relapse, infections and GVHD. The latter two constitute non-relapse mortality (NRM).^8,9^

Allo-HCT is a complex procedure, where patient outcome is not only determined by the disease but also depends on non-modifiable variables (comorbidities, donor availability) and modifiable ones, mainly depending on transplanting center decisions (donor choice, source of hematopoietic cells, type of conditioning regimen and GVHD prophylaxis).^5^

Therefore, systematic center-based monitoring of patient outcomes is essential to assess whether overall results align with expectations. Implementation of quality management systems was shown to be associated with improved overall outcomes, mostly in complex cases.^10,11^ Still, the time required to reach and maintain compliance with these systems may slow broader applications.^12^

The foundation for the accreditation of cellular therapy-joint accreditation committee of ISCT (JACIE) and the European society for blood and marrow transplantation (EBMT) have defined a set of outcome indicators intended to guide physicians in the systematic and periodic evaluation of patient outcomes as part of a self-assessment framework in allo-HCT.^13^

Although documented as less efficient than electronic data capture (EDC) software, spreadsheets are widely used for data management and processing in biomedical domains.^14,15^ Spreadsheets are also associated with unintentional errors such as incorrect cell references, formula corruption, and automatic conversion which have been documented to compromise the validity of research findings in various domains.^16^ This suggests that EDC-based registries and standardized analyses may reduce the risk of errors.

To date, however, while an EDC based European registry of allo-HCT collects longitudinal data, no standardized method exists to compute indicators and patient outcomes. While a well-designed benchmarking approach has enabled centers to compare their activity with that of peer institutions,^17^ it does not provide the annual epidemiological and outcome data required for local audits, and may lack scalability for high-frequency queries.

To tackle these limitations and use available data, an application was designed and developed which allows for automated analyses from local registries using formatted and non-formatted data^18^. AlloGraph is described here, a free and open-source interactive web-based dashboard which allows for fast, reproducible and interactive analyses of allo-HCT activities. It is already applied to the EBMT registry formatted dataset.

## METHODS

### Design of data processing and visualization

To account for possibly large panels, registry minimal data from 2018 to 2024 were retrieved from SFGM-TC participating centers on March 2025 using the EBMT Registry form: “Treatment overview report”. A de-identified sample of extracted data was provided to the application for test purposes. An example of the dataset format can be found at ‘data/test_sample.csv’ in the source code repository. Variables used are listed in **Supplementary Materials**.

### Data and user privacy

The app was designed to ensure that none of the provided data was stored in the app server. Data provided by the user are stored in the user web browser, namely client-side, and communicates with the Dash using ‘dcc.Storè. In AlloGraph, only the random-access memory (RAM) is used, meaning that all data is lost when upon page refreshing.

Neither the server nor the app use any cookie or connection survey, in order to ensure that user privacy is respected as well as compliance with European general data protection regulations.

### Data Pipeline

The application supports ‘CSV’ files or the provided import template. The application expects data conforming to the EBMT Registry “Treatment overview report” with standardized column names and formatted data without further modifications. None of the missing data are imputed. They are shown in the Dash representation to allow users identifying gaps in the dataset.

### Data analysis

Data were analyzed according to bone marrow-related standard data analysis.^19^ Categorical data are described as percentages and represented with bar plots. Continuous variables are summarized as median with interquartile range (IQR) or 95% confidence interval (CI) and boxplots. Events retained for GVHD- or relapse-free survival (GRFS) are acute or chronic GVHD with systemic immunosuppressive therapy, or relapse. Overall survival (OS) and GRFS analysis were performed with Log rank test and Kaplan-Meier representation. NRM was defined as death without previous relapse. Relapse, NRM and GVHD cumulative incidences were computed with Fine and Gray, with death as a competing event. Python libraries used were pandas, numpy, scipy, lifelines, plotly and datetime. Indicators were validated with the SFGM-TC scientific committee. Their definitions are detailed in ‘pages/indics.py’ file of AlloGraph repository and provided as plain text in **Supplementary Materials**.

### User survey

Once AlloGraph was deployed, users were asked to complete a survey hosted by REDCap Tours University. The survey asked users to describe their past experience of data analysis for local monitoring of activity, then a set of questions regarding user feedback.

### Source code

Source code of the software is available on an open repository (https://github.com/LaitEntier/AlloGraph) and long term preserved in Software Heritage (https://archive.softwareheritage.org/swh:1:dir:a3fc1be934aa36c057e520dc3429b2e14d2a250f).

### Ethical considerations

The app was developed with de-identified data and according to Helsinki declaration. The project was approved by the SFGM-TC board (DASHALLO project).

## RESULTS

### Program architecture

The design and main architecture elements of AlloGraph are described in **Figure 1**. Briefly, the app is hosted on a virtual machine on a secure server. On the virtual machine, the source code is pulled from the git repository then compiled locally and run on the webserver-side. The app relies on python and ‘dash’ library. The main entry point is ‘app.py’ which initializes the dash app providing dashboard activity survey. The codebase is organized with three main modules: reusable utility modules (‘modules/’), page-specific implementations (‘pages/’), and visualization components (‘visualizations/’). The source code hosted on a git repository was cloned on a Linux virtual machine hosted by Tours University server. The app is online on https://allograph.univ-tours.fr. Input data are provided by the user after downloading their local data from the European Registry or by conforming their own data with the provided import template in the home page.

**Figure 1.**
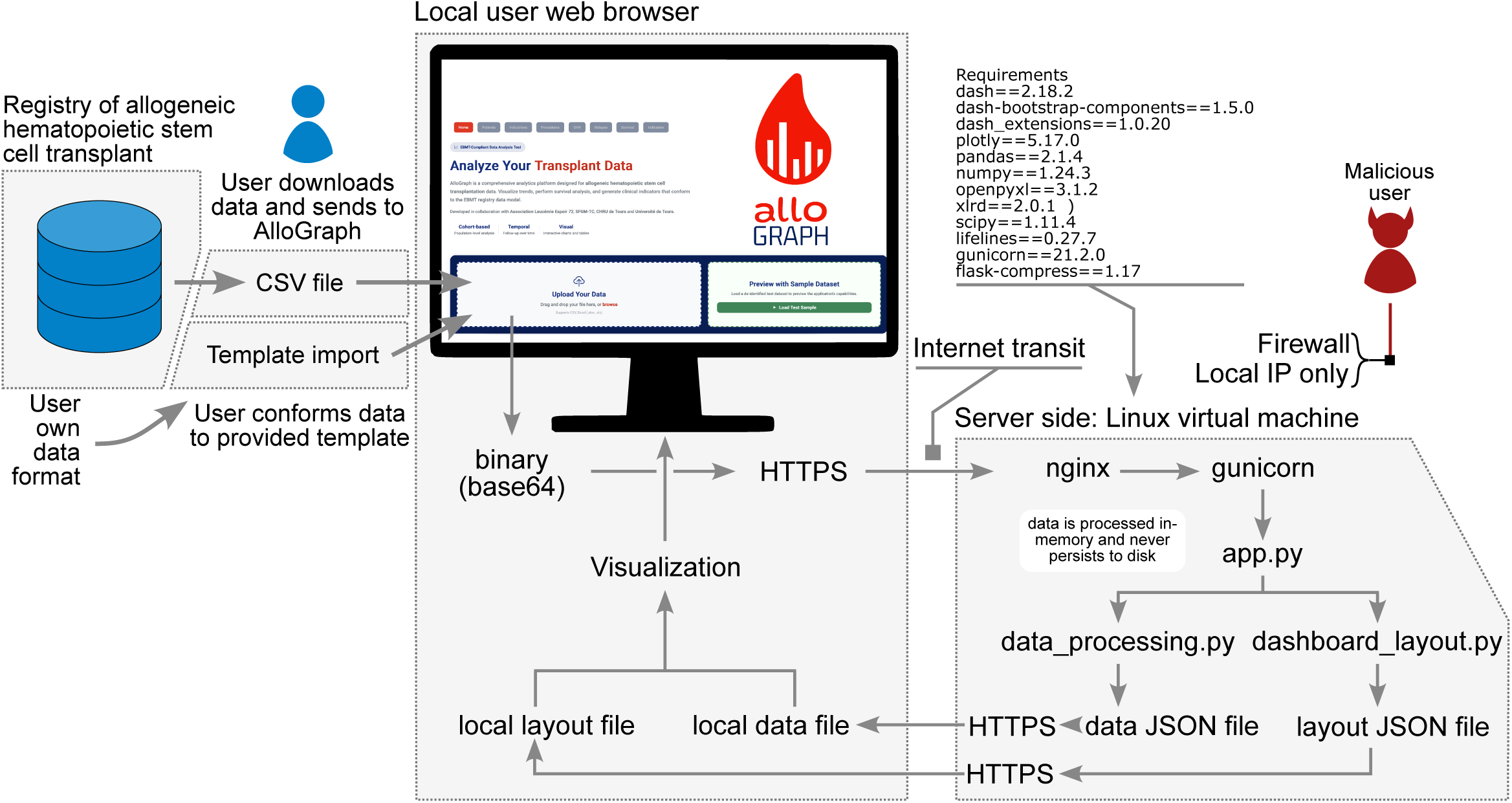
Architecture of AlloGraph.

The input file is directly converted to a binary (base64) format, then, after a secured transfer with a transport layer security (TLS) protocol, moved to the server side. TLS is the secured hypertext transfer protocol (HTTP) corresponding to HTTPS. It provides confidentiality with a secure data transmission channel associating cryptographic data encryption and required authentication from the user-side to the server-side. Upon arriving on the server-side, data are decrypted then stored in python memory buffer, in RAM. This implies that none of the data is written on the disk and the memory can be purged at anytime. All data manipulations stay in this memory buffer. As a consequence, the remote server does not host any data and access by malicious users would require a complex attack to access the RAM allocated to a virtual machine hosted by an authorized and local internet protocol (IP)-only server protected by firewalls.

Visualizations are the next step. Processed data and visualization are transformed to JSON files sent back to the user with the TLS protocol, allowing for visualization in the web browser. Once the user closes the web browser or selects the “void data” option, data is purged from the browser and server RAM memory. None of this data is written on the hard drive.

### Input data and dashboard visualizations

Optimal use of AlloGraph is provided by drag-n-dropping a CSV file formatted in the European allo-HCT registry. Otherwise, AlloGraph provides a data import template in which users can enter their own values. For evaluation purposes, AlloGraph provides a preview input which includes de-identified simulated data **(Figure 2A)**.

**Figure 2.**
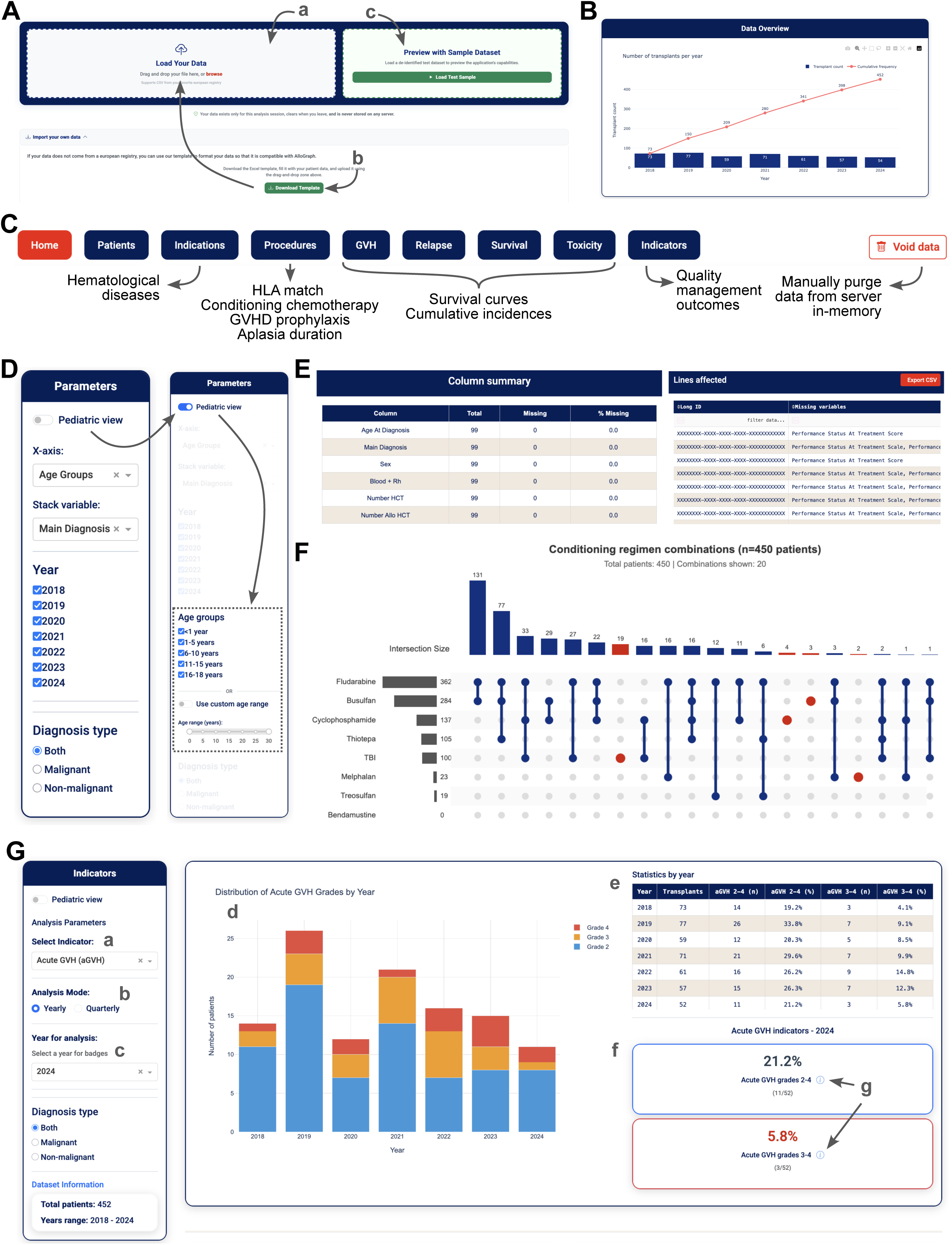
Overview of AlloGraph. A. Options for data input. User can either upload directly registry formatted data by drag-n-dropping the file or after choosing it with “browse” function (a). An import template is provided that the user must conform to (b). A preview dataset with simulated data is provided as an example of AlloGraph functionalities (c). B. The first plot shown provides an overview of the number of treatments performed yearly. C. The navigation bar shows the tabs with the corresponding data analysis and the “Void data” function to manually purge in-memory data. D. Navigation within each tab is allowed by a sidebar with options for plot drawing, axes change or filter/add a stack variable. Year selection and diagnosis type is provided to allow selected year analysis and choose malignant or non-malignant diseases. A pediatric view can be activated with a button to access additional filters to select children and adolescent young adults. E. At the bottom of each page, a table with the summary of missing data and a CSV export are provided F. Example of one of the visualizations provided. Here a plot shows the combinations of conditioning treatments. G. Quality management system indicators tab. Users choose the indicator (a) and a yearly or quarterly visualization (b) then a year of analysis (c). Visualization is provided at the center of the page (d) accompanied by a table with the corresponding values (e). Indicators are shown at the bottom right side of the page and the method used to determine indicator values are shown by hovering over the i symbol (g).

Once the file is provided, the first tab displays the annual number of allogeneic transplants and cumulative activity **(Figure 2B)**. A set of tabs is shown in the navigation bar allowing users to select the data to analyze **(Figure 2C)**. In each tab, using the “Parameters” sidebar, the user can customize the visualization by choosing the X-axis and stack variable to display. For pediatricians, a specific sidebar was designed to allow subgroup analyses according to specific age **(Figure 2D)**. At the bottom of the page a summary of studied and missing variables is provided along with specific missing values that can be exported in ‘CSV’ to complete original data **(Figure 2E)**.

The second tab describes patient characteristics, including age and performance status. The third tab presents disease indications by underlying diseases. The fourth tab details transplant procedures, including donor type, graft source, CMV donor/recipient status, conditioning intensity, individual and combined conditioning agents **(Figure 2F)**, individual and combined GVHD prophylaxis treatments, and aplasia duration. The fifth tab displays cumulative incidence curves for acute and chronic GVHD, with a filtering option by GVHD grade. The sixth tab presents cumulative incidence of relapse. The seventh tab shows overall survival and GRFS curves. Next, the eight tab describes toxicity as evaluated by NRM. The ninth tab displays JACIE quality indicators. Each indicator can be selected via a dropdown menu, and upon year selection, indicators are visualized by year and quarter, with graphical representation linked to numerical values. For each tab, a spreadsheet summarizes missing data **(Figure 2G)**.

### User experience survey

To evaluate the relevance of the app design and function, an online survey was organized between centers that had initially shared their activity data for AlloGraph development. Among the 45 participating centers, 30 respondents were recorded within 24 (53%) centers. Most respondents were physicians (n=21, 70%) and data managers (n=6, 20%), followed by clinical research assistants (n=3, 10%). A majority were from French centers (n=27, 90%) but also from Switzerland (Geneva, n=2, 7%) and one (3%) from Beyrouth in Lebanon. Treated patients were 24 adults (80%), 11 adolescent/young adults (AYA) (37%) and 12 children (40%) **(Table 1)**.

**Table 1.** Characteristics of survey respondents.

| **Respondent** | **n=30** |
| --- | --- |
| **Functions, n (%)** |  |
| Physician | 21 (70) |
| Data manager | 6 (20) |
| Clinical research assistant | 3 (10) |
| **Country, n (%)** |  |
| France | 27 (90) |
| Switzerland | 2 (7) |
| Lebanon | 1 (3) |
| **Age of treated patients in the center, n (%)** |  |
| Adults | 24 (80) |
| Adolescent young adults | 11 (37) |
| Children | 12 (40) |
| **Declared time of analysis, median (Q1, Q3)** | 2 (2, 9) days |
| Not answered, n | 10 |
| **Existing solution for analysis, n (%)** |  |
| Spreadsheet | 15 (68) |
| Automated program | 4 (18) |
| No solution | 2 (9) |
| Manual analysis | 1 (5.0) |
| Not answered | 7 |

Users were asked to describe their routine activity assessment practices. Median time required for analysis was estimated by 20 (67%) respondents at 2 (IQR: 2-9) days. Existing tools used for analysis were indicated by 22 respondents (73%) and were mostly spreadsheets (n=15; 68%), followed by automated program (n=4; 18%), manual analysis (n=1; 5%) while two (10%) declared that no specific tool was available **(Table 1)**.

To evaluate the applicability and consistency of AlloGraph regarding clinical practice and routinely assessed outcome, respondents were questioned on the app design. All respondents reported that the app was always and most of the time intuitive (n=19; 63% and n=11; 37%, respectively). Data were respectively reported as always (n=5; 17%), most of the time (n=20: 67%) and sometimes (n=4; 13%) displayed as expected. Respondents evaluated that navigation across the different tabs was always (n=9; 30%), most of the time (n=19; 63%) and sometimes (n=2; 7%) easy. AlloGraph was reported to respectively always (n=2; 7%), most of the time (n=10; 33%), sometimes (n=10, 33%), rarely (n=2; 7%) and never (n=1; 4%) bring a potential help in clinical and research decision making **(Figure 3A)**.

**Figure 3.**
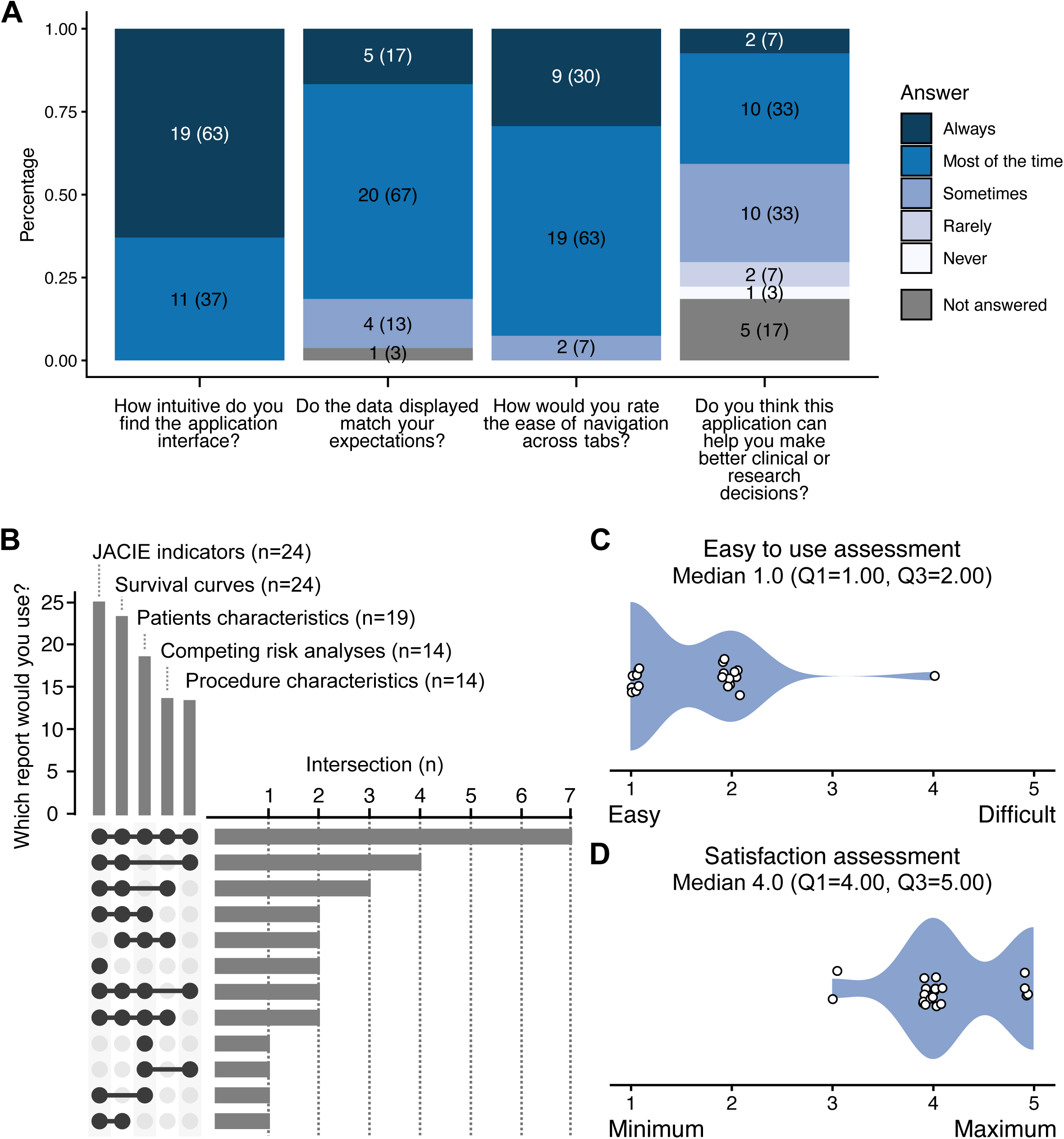
User experience survey. A. Stacked barplots showing answers regarding app design and clinical consistency B. Plot describing the reported most useful features of AlloGraph C. Violin plot depicting perceived ease of use D. Violin plot showing satisfaction levels

Respondents were then asked to describe which tab was the most useful for their survey activity. All respondents answered this question, the reported most useful reports were JACIE indicators (n=25; 83%), survival curves (n=24; 80%) and patient characteristics (n=19; 63%). In a combination analysis, 7 (23%) respondents reported that all tabs were useful and 4 (13%) reported that JACIE indicators, survival curves and procedure characteristics were useful **(Figure 3B)**.

To address overall scalability and consistency of the app, respondents were asked to evaluate whether AlloGraph was easy to use and state their satisfaction on a Likert scale. The app was reported to be easy to use for most respondents with a median score of 1.0 (Q1=1.0, Q3=2.0) **(Figure 3C)**. Overall satisfaction reached 4 to 5 for most users with a median score of 4.0 (Q1=4.0, Q3=5.0) **(Figure 3D).**

Technical difficulties using the app were reported by 6 respondents (21%): one had difficulty to visualize specific AYA patients, one reported difficulty relative to matching data not available in the input file, two reported that censoring information was not clearly shown, and one had trouble exporting data from the EBMT registry. To address these issues, in the current version of the app: (i) ticks for censored patients in survival curves and a risk table were implemented, (ii) an age subgroup selection tool with fixed and custom ranges to select, (iii) clarification on donor/recipients HLA matching were implemented, and (iv) a tutorial for data extraction is now provided.

Finally, the potential impact of AlloGraph on current analysis practices was evaluated. Twenty-five respondents (89%) answered that the app will change their way of monitoring their activity (2 did not answer this question). All 30 (100%) respondents would recommend AlloGraph to their peers.

## DISCUSSION

AlloGraph was designed and developed to allow for registry data analysis with a reproducible, standardized and easy-to-use tool, initially applicable to local monitoring of allo-HCT activity. High levels of usability, satisfaction, and usefulness were reported by clinicians and data managers.

In the survey, most respondents reported requiring a median of 2 days to perform routine activity assessments and predominantly (68%) relied on spreadsheet-based analyses. Healthcare systems are increasingly producing large volumes of real-world data in clinical registries, electronic health records, and administrative databases. Their routine use remains limited by the complexity of data extraction, analysis, and interpretation.^20,21^ This is particularly relevant in transplantation medicine, where extensive registry infrastructures have been developed such as the EBMT Registry which collects longitudinal data. Still, analysis is restricted to advanced analytical expertise which may vary substantially across centers.^17,22^ In addition, even if the use of spreadsheet seems easy, it requires a complex validation process to mitigate errors.^23,24^ Applications such as AlloGraph may provide significant opportunities for improving the accessibility of registry-based data and reduce analytical burden.^12^

AlloGraph is an open-source software that provides the code in a git-based repository, cloned in Software Heritage. This means that AlloGraph can be deployed anywhere with a protected.^25,26^ Consequently, potential local development branches may emerge in the future. To allow its use without expertise in informatics, AlloGraph was made available on an academic web-server. Accordingly, by providing automated computation of patient characteristics, donor type, conditioning regimen, survival analyses and activity indicators, AlloGraph enables physicians to explore their local activity without requiring programming expertise or dedicated bioinformatics support.

The concept of learning health systems emphasizes the continuous integration of clinical data into knowledge able to inform healthcare delivery and quality improvement.^27^ In this context, visualization platforms such as AlloGraph may represent an intermediary layer between data repositories and end users. It also integrates the goal of quality system improvement encouraged by the EBMT.^13^ Such democratization of access to healthcare data has been increasingly recognized as a key component of modern digital health infrastructures.^28^

All respondents considered AlloGraph to be intuitive, and, overall, satisfaction scores were high. The overall satisfaction level declared in the survey is consistent with previous successful implementation of healthcare dashboards.^29–31^

Interestingly, 89% of respondents anticipated that the application could modify their routine monitoring practices. These findings suggest that adoption barriers may be reduced when analytical tools are designed around existing clinical workflows rather than requiring users to adapt to complex software environment.

Concerns regarding confidentiality and compliance to regulation are the most significant barriers to the deployment of online digital health solutions.^32^ To address this limit, data transfer to the server is secured by a TLS protocol. The website is hosted on an academic server which is protected by firewalls and only allows connections with authorized IP address. AlloGraph is based on exclusively RAM usage without persistent storage. Data are thus automatically removed from server memory and the browser environment at the end of the user session, avoiding the risk of personal and health data leaking. This privacy-by-design approach aligns with contemporary recommendations for secure handling of health data and may facilitate its implementation in environments where institutional or regulatory constraints limit the transfer of sensitive clinical information.^32,33^ This development strategy may serve as a standard for clinical dashboard developments.

Respondents identified JACIE indicators, survival analyses, and patient characteristics as the most valuable tabs. This may be explained by the fact that indicators are an essential part of center evaluations.^12,17,34^ Furthermore, dashboards have been shown to be most effective by focusing on meaningful visualizations which support clinical or organizational decisions.^29,30^

Some limitations should be acknowledged. The study relied primarily on subjective measures of usability and satisfaction. To date, dashboards have not been shown to impact clinical decision or physician behavior in prescriptions^35^. Objective assessments of efficiency, accuracy, or clinical impact will be of interest to explore how such tools as AlloGraph may impact healthcare systems and clinical practices. Still, considering that AlloGraph is not a research program and that clinical decision-making should be based on expert statistical analysis, it was chosen here not to implement comparisons between patient categories, to avoid spurious conclusions based on data unadjusted for potential multiple biases. Also, the sustainability and maintenance of the website is a threat to any dashboard.

Altogether, this suggests that AlloGraph addresses an unmet need for accessible and secure visualization of transplant activity data. By encouraging centers to use registry data or routinely collected data to monitor their own activity, the implementation of AlloGraph may therefore lead to measurable improvements in quality of registry data, number of missing data, accreditation processes, research productivity, even in a time when centers are short on medical time. This will require a specific study in the future.

In conclusion, by combining secure in-memory processing with intuitive visualization tools, the AlloGraph platform addresses important barriers to the routine exploitation of real-world data. This suggests that such platforms may help bridge the gap between data collection and data-driven decision-making. Beyond its application to allo-HCT, AlloGraph illustrates how registry-derived data can be transformed into actionable information using a user-centered digital health approach.

## Data Availability

Data and source code of the software are available on an open repository (https://github.com/LaitEntier/AlloGraph) and long term preserved in Software Heritage (https://archive.softwareheritage.org/swh:1:dir:a3fc1be934aa36c057e520dc3429b2e14d2a250f).

## Acknowledgments

Authors would like to (i) express their sincere gratitude to *Tours University DSI* for their support in hosting the app and (ii) address their profound thankfulness to EBMT President: Prof. Ibrahim Yakoub-Agha and to IT and Registry Committee for insightful discussions following the app deployment: Annelot van Amerongen, Alvaro Ruiz Hernandez, Igor Ariz, Ignacio Garcia and Hendrikus Bouwman. Medical writing for this manuscript was assisted by MPIYP (MC Béné), Paris, France. Graphic identity was conceptualized and performed with the help of Lucie Clarysse (https://comsci.art/). The software is open source and has been registered at the “Agence pour la Protection des Programmes”: https://secure2.iddn.org/app.server/certificate/?sn=2026090021000&key=d8604eb9c3b91734378da292007e17865c6877f7de4a0420dea7daf23dfaba12&lang=fr

## Authors contributions

NV was responsible for project administration and secured funding to the project.

NV and AC conceptualized the app and designed the methodology and data visualization.

AC developed the source code and deployed AlloGraph

NV, DM, VL, AV, ME, EG and AC performed first beta tests of the app.

NV and AC wrote the original draft of the manuscript.

All authors critically reviewed and accepted the manuscript.

## Data Sharing Statement

Not applicable

