## Supplementary Methods for "AlloGraph: open-source web-based scalable platform for registry-driven monitoring of allogeneic hematopoietic cell transplant activity"

Variables retrieved from registry minimal data

Long ID, Short ID, Promise ID, Sex, Date Of Birth, Blood Group, Rhesus Factor, Initials First Name, Initials Last Name, Date Diagnosis, Main Diagnosis, Subclass Diagnosis, Treatment CIC, Treatment Type, Treatment Date, Number HCT, Number Allo HCT, Performance Status At Treatment Scale, Performance Status At Treatment Score, Disease Status At Treatment, CMV Status Donor, CMV Status Patient, Donor Type, Source Stem Cells, Source Stem Cells 2, Match Type, Conditioning Regimen Type, Prep Regimen Bendamustine, Prep Regimen Busulfan, Prep Regimen Cyclophosphamide, Prep Regimen Fludarabine, Prep Regimen Melphalan, Prep Regimen Thiotepa, Prep Regimen Treosulfan, Prophylaxis, Prophylaxis Drug 1, Prophylaxis Drug 2, Prophylaxis Drug 3, Prophylaxis Drug 4, Prophylaxis Drug 5, Prophylaxis Drug 6, TBI, TBI Dose Gray, Date Of Last Follow Up, First aGvHD Maximum Score, First Agvhd Occurrence, First Agvhd Occurrence Date, First cGvHD Maximum NIH Score, First Cgvhd Occurrence, First Cgvhd Occurrence Date, First Relapse, First Relapse Date, First Best Response, First Best Response Date, Platelet Reconstitution, Date Platelet Reconstitution, Anc Recovery, Date Anc Recovery, Date Subsequent Treatment, Performance Scale At Last FU, Performance Score At Last FU, Cgvhd Maximum Nih Score At Last Fu, Cgvhd Occurrence At Last Fu, Status Last Follow Up, Death Cause, Death Date

Annual indicators definitions

**Acute Graft-versus-Host Disease (aGVH)**
Function: process_gvha_data()
Variables: First Agvhd Occurrence Date, Treatment Date, First aGvHD Maximum Score
Method: Time to aGVH = First Agvhd Occurrence Date - Treatment Date (days)
Inclusion : time ≤ 100 days AND Grade ∈ {Grade 2, Grade 3, Grade 4}

GvHa (%) = (Number of cases / Total number of transplant) × 100

**Chronic Graft-versus-Host Disease (cGVH)**
Function: process_gvhc_data()
Variables: First Cgvhd Occurrence Date, Treatment Date, First cGvHD Maximum NIH Score
Method: Time to cGVH = First Cgvhd Occurrence Date - Treatment Date (in days)
Inclusion: Time within 365 days AND Score ∈ {Mild, Moderate, Severe}
Transformation
- Limited → Mild
- Extensive → Severe

GvHc (%) = (Number of cases / Total number of transplant) × 100

**Non-Related Mortality (NRM or toxicity related mortality)**
Function: process_trm_data()
Variables: Status Last Follow Up, Death Cause, Date Of Last Follow Up, Treatment Date
Method: Follow-up days = Date Of Last Follow Up - Treatment Date

Inclusion filter :
- Death Cause ∈ {'Cellular therapy-related cause of death', 'HCT-related cause of death'}
- AND (Status = 'Dead' OU (Status = 'Alive' ET Follow-up days ≥ 365))

NRM D30 = NRM event AND Follow-up days ≤ 30 days
NRM D100 = NRM event AND Follow-up days ≤ 100 days
NRM D365 = NRM event AND Follow-up days ≤ 365 days

NRM (%) = (Number of NRM events / Total number of transplants) × 100

**Overall Survival**
Function: process_survival_data()
Variables: Status Last Follow Up, Date Of Last Follow Up, Treatment Date
Method: follow up days : Date Of Last Follow Up - Treatment Date
Inclusion filter:
- Status ∈ {'Dead', 'Died after conditioning but before main treatment'}
- OR (Status = 'Alive' AND Follow-up days ≥ 365)

Death D30 = death AND Follow-up days ≤ 30
Death D100 = death AND Follow-up days ≤ 100
Death D365 = death AND Follow-up days ≤ 365

Survival (%) = (Events / Total number of transplant) × 100

**Transplantation Success / Platelet Reconstitution**
Fonction : process_prise_greffe_data()
Variables: Date Platelet Reconstitution, Treatment Date
Méthode de calcul : Time reconstitution = Date Platelet Reconstitution - Treatment Date (days)
Inclusion: non-missing data AND Délai ≤ 100 jours
Success (%) = (Success / Total number of valid data) × 100

**ANC Recovery**
Function: process_sortie_aplasie_data()
Variables: Date ANC Recovery, Treatment Date
Method: Time to ANC recovery = Date ANC Recovery - Treatment Date (days)
Inclusion filter:
- Non-missing data AND Recovery delay <= 28 days

Success = Number of patients with ANC recovery <= 28 days
ANC Recovery Rate (%) = (Success / Total number of valid data) x 100

**Relapse**
Function: process_rechute_data()
Variables: First Relapse, First Relapse Date, Treatment Date
Method: Time to relapse = First Relapse Date - Treatment Date (days)
Inclusion filter:
- First Relapse = "Yes" AND First Relapse Date non-missing

Relapse D100 = First Relapse = "Yes" AND Time to relapse <= 100 days
Relapse D365 = First Relapse = "Yes" AND Time to relapse <= 365 days

Relapse Rate (%) = (Number of relapses / Total number of valid data) x 100
Visualization: Curve with datapoints at D100, D365.
